# Accurate genome-wide germline profiling from decade-old archival tissue DNA reveals the contribution of common variants to precancer disease outcome

**DOI:** 10.1101/2022.03.31.22273116

**Authors:** Daniela Nachmanson, Meghana Pagadala, Joseph Steward, Callie Cheung, Lauryn Keeler Bruce, Nicole Q. Lee, Thomas J. O’Keefe, Grace Y. Lin, Farnaz Hasteh, Gerald P. Morris, Hannah Carter, Olivier Harismendy

## Abstract

**Background:** Inherited variants have been shown to contribute to cancer risk, disease progression, and response to treatment. Such studies are, however, arduous to conduct, requiring large sample sizes, cohorts or families, and more importantly, a long follow-up to measure a relevant outcome such as disease onset or progression. Unless collected for a dedicated study, germline DNA from blood or saliva are typically not available retrospectively, in contrast to surgical tissue specimens which are systematically archived.

**Results:** We evaluated the feasibility of using DNA extracted from low amounts of fixed-formalin paraffin-embedded (FFPE) tumor tissue to obtain accurate germline genetic profiles. Using matching blood and archival tissue DNA from 10 individuals, we benchmarked low-coverage whole-genome sequencing (lc-WGS) combined with genotype imputation and measured genome-wide concordance of genotypes, polygenic risk scores (PRS), and HLA haplotypes. Concordance between blood and tissue was high (r^2^>0.94) for common genome-wide single nucleotide polymorphisms (SNPs) and across 22 disease-related PRS (mean r=0.93). HLA haplotypes imputed from tissue DNA were 96.7% (Class I genes) and 82.5% (Class II genes) concordant with deep targeted sequencing of HLA from blood DNA. Using the validated methodology, we estimated breast cancer PRS in 36 patients diagnosed with breast ductal carcinoma in situ (11.7 years median follow-up time) including 22 who were diagnosed with breast cancer subsequent event (BSCE). PRS was significantly associated with BCSE (HR=2.5, 95%CI: 1.4–4.5) and the top decile patients were modeled to have a 24% chance of BCSE at 10 years, hence suggesting the addition of PRS could improve prognostic models which are currently inadequate.

**Conclusions:** The abundance and broad availability of archival tissue specimens in oncology clinics, paired with the effectiveness of germline profiling using lc-WGS and imputation, represents an alternative cost and resource-effective alternative in the design of long-term disease progression studies.

## Introduction

The study of the contribution of germline genetic variation to disease risk or treatment outcome typically requires blood or saliva samples as a source of constitutive DNA. Depending on the phenotype studied, such samples may not be banked and readily available. Samples may have to be prospectively collected, which hinders studies requiring long-term follow-up or obtained after contacting potential subjects of interest, which can be logistically and ethically challenging or impossible if a patient has relocated or died. In cancer research, there is a growing interest in directly profiling tumor tissue to obtain germline measures such as ancestry, polygenic risk, and HLA-typing [1]. Array-based genotyping followed by imputation from a reference population has been a standard method to genotype genome-wide SNPs in the human genome, but its compatibility with DNA obtained from archival tissue specimens remains to be established [2,3]. The approach can be challenging when the amount of tissue available for research is limited, which is often the case with surgical excisions of premalignant lesions or with most needle biopsies.

Recently low-coverage whole-genome sequencing (lc-WGS) has emerged as an attractive alternative to single nucleotide polymorphism (SNP) array by offering higher throughput at a reduced cost, reduced DNA input, and improved genotyping accuracy [4–6]. In fact, recent studies have shown the feasibility of using frozen tissue for germline profiling by imputing genotypes from off-target reads repurposed from tumor-targeted panel sequencing data, effectively equivalent to ultra-low coverage (less than 0.1x) whole-genome sequencing [1]. It is therefore likely that, in the absence of available targeted sequencing data, lc-WGS can be performed with DNA of lower quality and quantity to enable the imputation of germline variants from archival tissue specimens.

If accurate, such an approach could have important implications for the study of the contribution of inherited risk factors to the progression of pre-malignant disease. For many cancer types, the widespread adoption of cancer screening has led to an increase in the detection of pre-malignant lesions. Despite such efforts, screening has had limited impact on overall survival [7]. Clinical guidelines vary widely from watchful waiting or biopsy as for prostatic intraepithelial neoplasia to surgery and adjuvant treatment as for ductal carcinoma in situ (DCIS) of the breast [8,9]. In absence of reliable progression risk biomarkers and models, these interventions may have deleterious consequences at the two clinical extremes: delay in life-saving treatment or complications from overtreatment. DCIS is the most common breast cancer-related diagnosis, comprising ∼20% of annual cases in the U.S. [10]. In breast disease, factors that impact the risk of breast cancer subsequent event (BCSE), defined as an in situ or invasive breast cancer neoplasm developed at least 6 months after treatment of a DCIS diagnosis, include age, size, grade of the lesion, hormone receptor status, and molecular profile. Their combined effect in risk models such as the University of Southern California / Van Nuys Prognostic Index has not resulted in any reliable BCSE risk prediction model and additional, more in-depth molecular and histological characterization is needed [11–16].

Given the independence between DCIS and associated BCSE in upwards of 20% of cases as evidenced by molecular studies comparing genomic profiles of initial DCIS and subsequent ipsilateral BCSE, systemic risk factors need to be considered in addition to those related to the index lesion [17]. While penetrant germline pathogenic variants exist and represent strong risk factors in breast cancer susceptibility genes such as *BRCA1, BRCA2, CHEK2, PALB2*, and *PMS2*, they are only present in 1.5% of all women [18]. Meanwhile, population-based genome-wide association studies (GWAS), have identified multiple common variants associated with lifetime risk of invasive breast cancer (IBC) [2,19]. The same SNPs have also been associated with risk of DCIS demonstrating the shared genetic susceptibility for IBC and DCIS [20]. It is however unclear if these SNPs are also associated with DCIS progression. Polygenic risk scores (PRS) derived from the allelic burden of risk-associated SNPs are now being added to common breast cancer risk models, significantly improving their performance, with individuals in the top percentile having a 3-5 fold increase in lifetime risk relative to women with risk scores in the middle quintile of those studied [21,22]. It is thus possible that DCIS patients with elevated breast cancer PRS are also at higher risk of BCSE and the addition of PRS could improve DCIS prognostic models akin to lifetime breast cancer risk models. Since BCSE can occur years after the initial DCIS diagnosis and is uncommon - observed in 10 to 25% of patients after 10 years, depending on treatment and known risk factors - a retrospective study is much more feasible for the purposes of validation [23,24]. Formalin-fixed paraffin-embedded (FFPE) tissue (referred to as archival tissue) from the DCIS biopsy or resection are therefore the only source of genetic material available and their validity for genome-wide genotyping of germline variants would be critical to the feasibility of such study.

Here we evaluate the validity of repurposing archival tissue specimens for germline genetic studies. We performed lc-WGS and imputed genotypes for 10 pairs of matching blood and tumor tissue samples to benchmark the accuracy for calling genome-wide genotypes, HLA haplotypes, and for implementing PRS. The reported results indicate the high accuracy of germline genotypes and haplotypes obtained from archival tissue DNA. Using this methodology we estimate breast cancer PRS in 36 DCIS patients and demonstrate its association with BCSE.

## Results

### Concordance of lc-WGS imputed genotypes between blood DNA and FFPE tissue DNA

In order to establish the analytical validity of FFPE tissue DNA for germline genotyping and genotype imputation from lc-WGS, we selected 10 subjects including from European, African, and Asian ancestries with matching FFPE tissue and whole blood. The archival tissue blocks were between 3 and 9 years old and yielded between 5 and 176 ng of DNA, which was then prepared for sequencing with a low-input protocol (see Methods). Mean coverage depth was 0.92x (range 0.68-1.41x) and 0.7x (range 0.44-0.97) for blood and tissue, respectively. Genotypes were imputed using a Gibbs sampling method specifically designed for lc-WGS, which leverages haplotype reference panel information (1000G 30x NYGC reference panel - N=3,202 individuals; see Methods) [5,25]. Overall genotypes were imputed for 61,715,567 SNPs in each of the 20 samples, of which 43,274,690 (70.1%) were considered high quality (Impute INFO score >0.80) [26]. Genotype concordance between blood and tissue increased with the minor allele frequency (MAF) of the variant in the global population. For SNPs with MAF of 0.1 or more, the aggregate r^2^ was greater than 91% for all SNPs, and greater than 94% for high-quality SNPs (Figure 1a). The concordance between blood and tissue was not lower for the two individuals who were non-white (Figure S1). In contrast, the concordance was lower (87% at SNPs with MAF greater or equal to 0.1) when the sequencing coverage depth of the tissue DNA was lower (Figure S2). Overall, the strongest discordance between blood and tissue was observed for SNPs at MAF lower than 0.01 which are typically imputed with decreased accuracy irrespective of the sample type [27].

**Figure 1.**
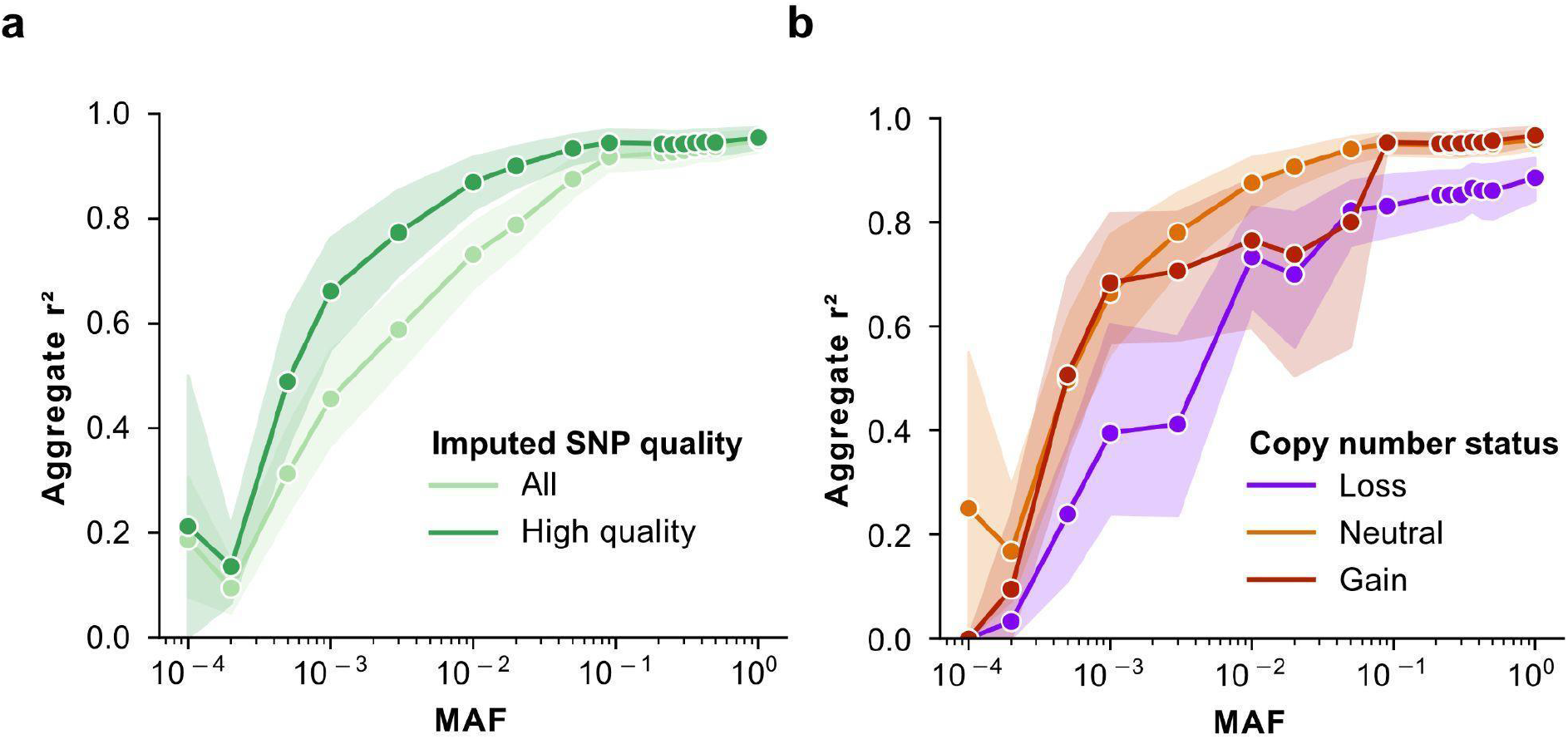
Assessment of genome-wide concordance of lc-WGS imputed genotypes in tissue versus blood of N=10 patients. **(a**,**b)** Genome-wide concordance (Pearson correlation coefficient squared - y-axis) of allele dosages across all genotyped SNPs between blood and tissue as a function of their minor allele frequency (MAF, x-axis). Concordance was calculated for each individual and each filtering category including genotype imputation quality (a) with all genotypes shown in light green and high-quality genotypes (INFO>80) in dark green, and copy number status of high-quality genotypes in tissue (b), from SNPs located in a region that was copy neutral (orange), gain (red) or loss (blue). For any given bin corresponding to a patient, MAF and filtering category had to have a minimum of 1,000 SNPs to be included. Error estimates from 95% confidence intervals computed from 1,000 bootstrapping iterations are indicated as shaded areas.

The presence of somatic mutations and copy number alterations (CNA) in DNA from malignant cells has the potential to decrease local imputation accuracy. In particular, CNA may play a larger role than somatic mutations, as recently reported [1]. We estimated the effect of CNA status on the genotype concordance between blood and tissue across SNPs located in DNA regions that are copy neutral, in a copy gain, or in a copy loss. The studied DNA samples had, on average, 15% of the genome (range 0 to 65%) involved in CNA while no CNA was detected in the blood (see Methods, Table S1). Common SNPs (MAF≥0.1) located in copy neutral or copy gain regions had a remarkable blood-tissue genotype concordance r^2^ higher than 95%, while those in regions of copy number loss showed lower concordance r^2^ of 83% (Figure 1b). The decreased imputation accuracy in areas of copy number loss can likely be explained by the decrease in allele-specific coverage depth, resulting in missed heterozygotes or a sparser scaffold for imputation.

We conclude that tissue-derived genome-wide genotypes faithfully represent germline profiles obtained from blood, especially at SNPs frequent in the population (MAF≥0.1). Discrepancies between tissue and blood can be explained by decreased coverage depth caused by technical (insufficient sequencing) or genetic (copy number loss) limitations and mainly affecting rare SNPs (MAF<0.01). These lower frequency SNPs are less likely to reach statistical significance in GWAS studies unless they have extreme effect size and therefore are rarely incorporated into PRS models. Taken together, the results suggest the feasibility of using archival tissues as a source of constitutive DNA in genetic studies relying on common SNPs.

### Concordance of tissue-derived PRS

We next sought to further validate the performance of tissue-based genome-wide genotyping to accurately estimate PRS in individuals. Germline variants can be used to estimate disease risk in individuals by summing the effects of previously identified risk alleles carried by an individual into a personalized PRS. The clinical utility of PRS is currently being evaluated in multiple settings, including breast cancer screening and surveillance, where elevated PRS can be included in lifetime risk models [28]. The ability to accurately estimate PRS retrospectively, using archival tissue DNA, would greatly improve the ability to conduct large retrospective studies with long-term outcomes. We investigated multiple PRS derived from GWAS of susceptibility to 16 cancer types, and 6 non-cancer phenotypes [29–31]. We computed a tissue and blood-derived PRS for 10 individuals (see Methods) using the imputed genotypes from lc-WGS sequencing data described above. Overall 93% (2,744 of 2,962) of PRS single-nucleotide variant sites were successfully imputed, 84% of which were high quality (Table S2). In each of the 16 cancer types, the tissue-derived PRS closely matched the blood-derived PRS, evidenced by high correlation coefficients (r≥0.9) in 12/16 of the PRS (Figure 2a). We saw similar results when evaluating PRS for non-cancer phenotypes, with 4/6 being highly correlated (r≥0.9) (Figure 2b). Differences between PRS in blood and tissue were associated with decreased tumor genome coverage (r=-0.26, p=0.02), but not with copy number loss (Figure S3). Overall we report that archival tissue DNA profiled with lc-WGS resulted in a reliable PRS estimate in an individual and preserved relative ranks in a cohort enabling studies such as the use case presented below.

**Figure 2.**
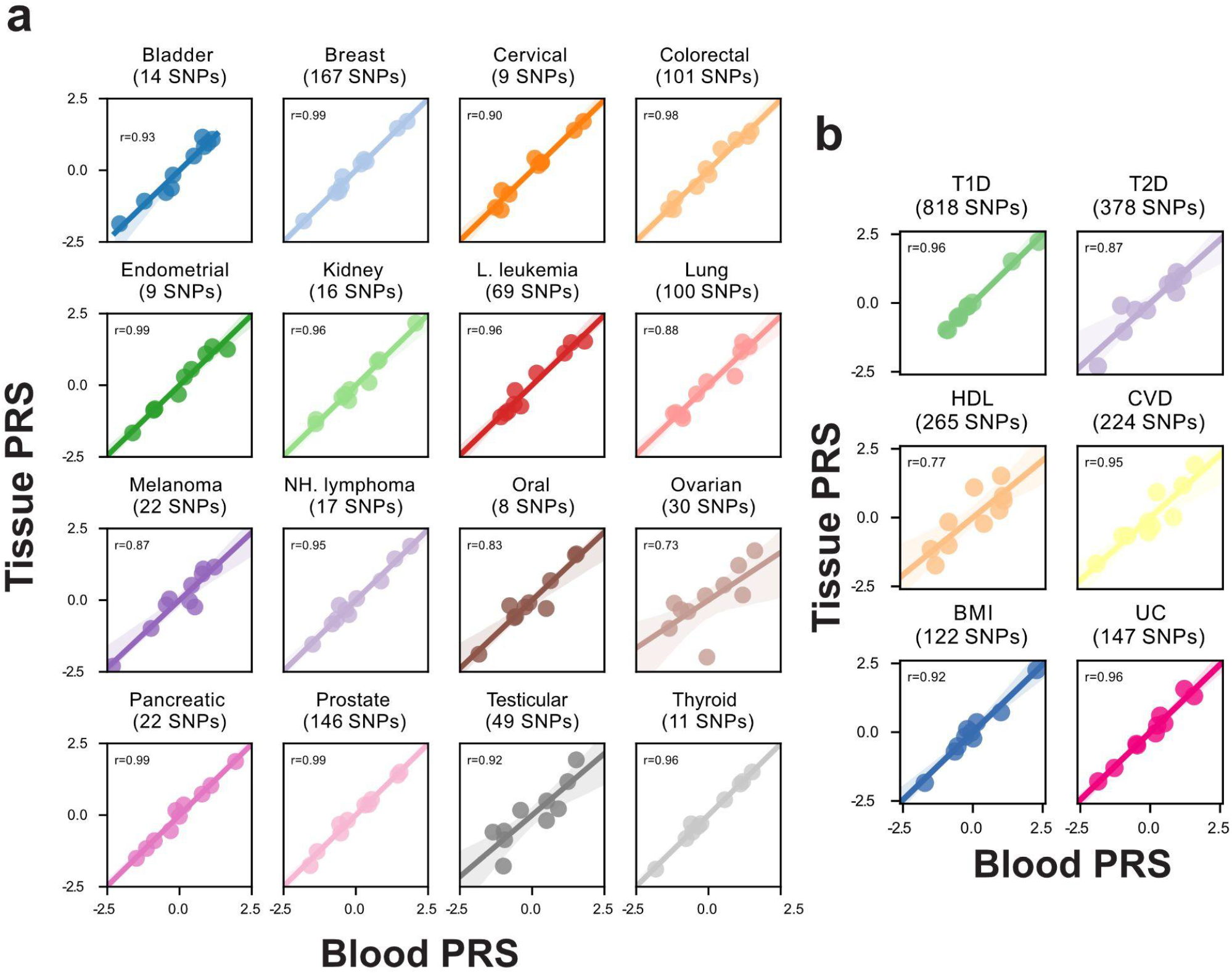
Blood versus tissue-derived PRS. **(a)** Cancer and **(b)** non-cancer PRS computed from imputed genotypes from lc-WGS of blood (x-axis) and tissue (y-axis) of the same patient. Spearman correlation coefficient, r, was measured between blood and tissue PRS values across N=10 patients, for each normalized PRS. T1D: Type 1 diabetes, T2D: Type 2 diabetes, HDL: High-density lipoprotein, CVD: Cardiovascular disease, BMI: Body mass index, UC: Ulcerative colitis.

### Contribution of breast polygenic risk scores to DCIS prognosis

We next demonstrated the utility of lc-WGS to investigate the contribution of breast cancer PRS to predict breast cancer subsequent events (BCSE - in situ or invasive, irrespective of laterality) after a DCIS diagnosis using a retrospective study design (Figure 3a). We assembled a cohort of patients diagnosed with pure DCIS (N=25 cases) who were then diagnosed with a BCSE at least 6 months after the DCIS diagnosis. We then complemented this cohort with a set of patients (N=25 controls) diagnosed with pure DCIS who did not develop a BCSE for at least 5 years. A median of 51.2 ng (range 6.6-300) of DNA was extracted from the primary DCIS FFPE specimen archived between 6 and 25 years (Table S3). The extracted DNA was sequenced to an average coverage depth of 0.89x (range 0.2-1.8x) (see Methods). Fourteen out of 50 (28%) samples yielded insufficient coverage (N=5) or had evidence of contamination with another patient (N=9) and were excluded, leaving 22 cases and 14 controls for analysis (Table 1). The median time to BCSE was 6.2 years (min: 1.4, max: 10.9), and patients without BCSE had a median time to follow-up of 11.7 years (min: 6.7, max: 19.6). Cases and controls were approximately matched for age, ancestry, DCIS size, grade, and ER status (Table S4). We then performed imputation as described earlier which resulted in high-quality genotypes at a total of 27,605,021 SNP loci.

**Table 1.** Clinical characteristics of the DCIS cohort.

| **Patient** | **BCSE?** | **Race** | **Ethnicity** | **Age range at diagnosis** | **Type of surgery** | **Pathologic size (cm)** | **Nuclear grade** | **ER Status (0/1)** | **Days to BCSE or last followup** |
| --- | --- | --- | --- | --- | --- | --- | --- | --- | --- |
| OXPA003 | Yes | White | Non-Hispanic | 41-50 | Lumpectomy | 0.8 | 1 | + | 2554 |
| OXPA028 | Yes | White | Non-Hispanic | 51-60 | Mastectomy | 0.5 | 1 | + | 1012 |
| OXPA033 | Yes | White | Non-Hispanic | 71-80 | Lumpectomy | NA | 1 | + | 1024 |
| OXPA036 | Yes | White | Non-Hispanic | 51-60 | Lumpectomy | NA | 1 | + | 2296 |
| OXPA161 | Yes | White | Non-Hispanic | 61-70 | Mastectomy | 0.4 | 1 | + | 3752 |
| OXPA166 | Yes | White | Non-Hispanic | 51-60 | Lumpectomy | 0.7 | 1 | + | 7037 |
| OXPA020 | Yes | White | Non-Hispanic | 51-60 | Lumpectomy | NA | 1 | NA | 5967 |
| OXPA021 | Yes | White | Non-Hispanic | 41-50 | Lumpectomy | 0.9 | 1 | NA | 2398 |
| OXPA527 | Yes | White | Hispanic | 41-50 | Lumpectomy | 0.7 | 1 | NA | 2203 |
| OXPA002 | Yes | Asian | Non-Hispanic | 41-50 | Lumpectomy | 1.4 | 2 | + | 2492 |
| OXPA006 | Yes | White | Non-Hispanic | 61-70 | Lumpectomy | 1 | 2 | + | 3022 |
| OXPA032 | Yes | Asian | Non-Hispanic | 41-50 | Lumpectomy | 2.5 | 2 | + | 502 |
| OXPA044 | Yes | White | Non-Hispanic | 51-60 | Lumpectomy | 0.9 | 2 | + | 2147 |
| OXPA064 | Yes | White | Non-Hispanic | 71-80 | Lumpectomy | 0.4 | 2 | + | 553 |
| OXPA179 | Yes | White | Hispanic | 71-80 | Lumpectomy | 1.2 | 2 | NA | 3077 |
| OXPA147 | Yes | White | Non-Hispanic | 61-70 | Lumpectomy | 1.1 | 3 | + | 1981 |
| OXPA185 | Yes | White | Non-Hispanic | 41-50 | Lumpectomy | 0.3 | 3 | + | 497 |
| OXPA150 | Yes | White | Non-Hispanic | 51-60 | Lumpectomy | 0.4 | NA | + | 2402 |
| OXPA153 | Yes | White | Non-Hispanic | 61-70 | Lumpectomy | 0.5 | NA | + | 3970 |
| OXPA246 | Yes | White | Non-Hispanic | 71-80 | Lumpectomy | 0.5 | NA | + | 1179 |
| OXPA267 | Yes | White | Non-Hispanic | 61-70 | Mastectomy | NA | NA | + | 3083 |
| OXPA151 | Yes | White | Non-Hispanic | 71-80 | Lumpectomy | NA | NA | NA | 1029 |
| OXPA644 | No | White | Non-Hispanic | 51-60 | Lumpectomy | 1.1 | 1 | - | 2456 |
| OXPA347 | No | White | Non-Hispanic | 81-90 | Lumpectomy | 5 | 1 | + | 3894 |
| OXPA508 | No | White | Non-Hispanic | 51-60 | Lumpectomy | 0.9 | 1 | + | 2570 |
| OXPA172 | No | White | Non-Hispanic | 71-80 | Lumpectomy | 1.8 | 1 | NA | 6903 |
| OXPA295 | No | White | Non-Hispanic | 51-60 | Lumpectomy | 1.2 | 1 | NA | 4903 |
| OXPA092 | No | White | Non-Hispanic | 51-60 | Lumpectomy | 0.5 | 2 | + | 3822 |
| OXPA156 | No | White | Hispanic | 41-50 | Lumpectomy | 0.5 | 2 | + | 7170 |
| OXPA392 | No | White | Non-Hispanic | 51-60 | Lumpectomy | 0.6 | 2 | + | 3709 |
| OXPA445 | No | White | Non-Hispanic | 51-60 | Lumpectomy | 1.3 | 2 | + | 3594 |
| OXPA501 | No | Asian | Non-Hispanic | 41-50 | Lumpectomy | 1.2 | 2 | + | 3044 |
| OXPA530 | No | White | Hispanic | 61-70 | Lumpectomy | 1.5 | 2 | + | 3167 |
| OXPA540 | No | Asian | Non-Hispanic | 41-50 | Lumpectomy | 1.8 | 2 | + | 2941 |
| OXPA182 | No | White | Non-Hispanic | 51-60 | Lumpectomy | 0.5 | 3 | NA | 4543 |
| OXPA146 | No | White | Non-Hispanic | 41-50 | Lumpectomy | 1 | NA | + | 7067 |

**Figure 3.**
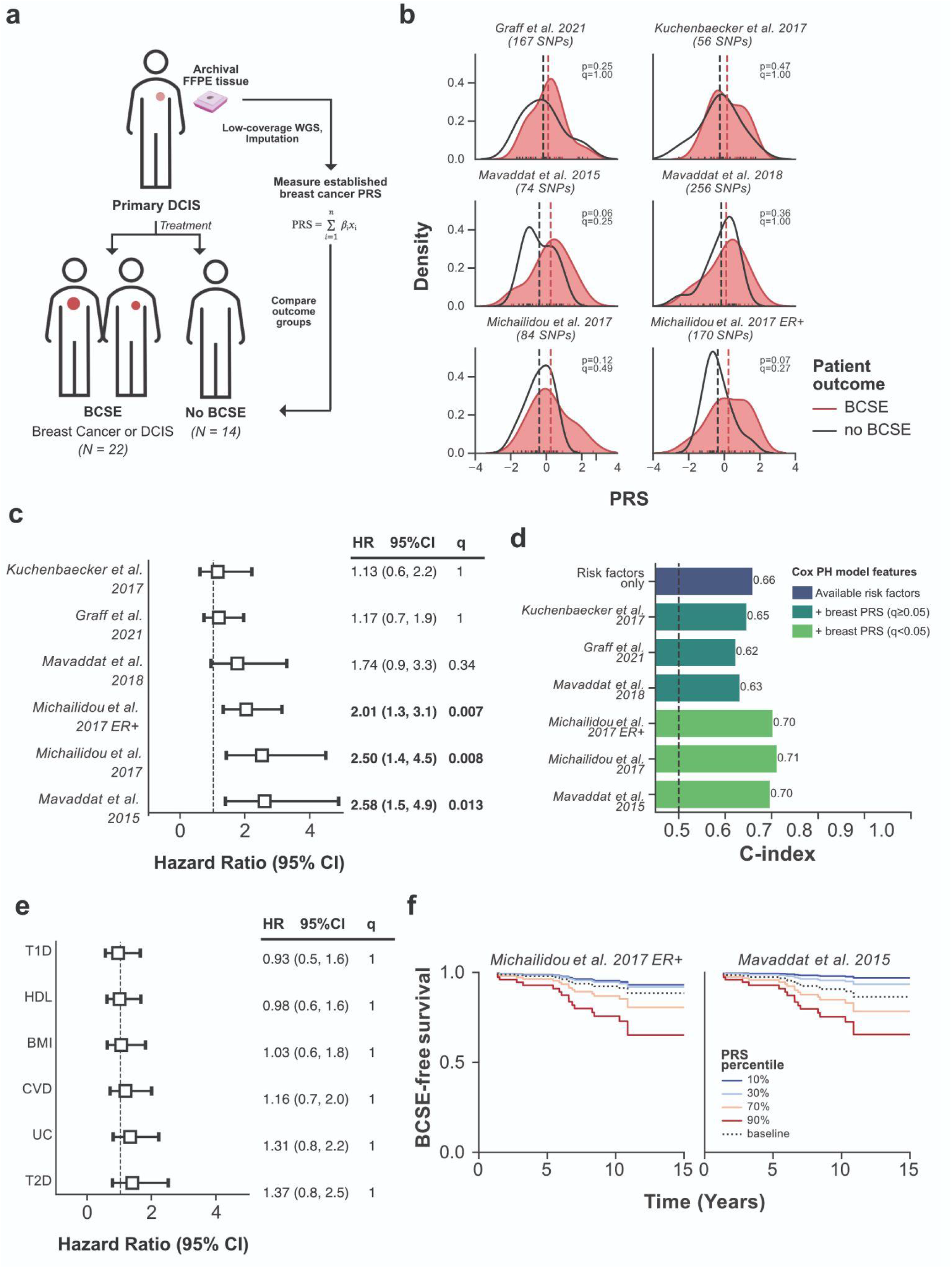
Breast cancer polygenic risk score in DCIS patients with and without a breast cancer subsequent event. **(a)** Schematic overview of the study design. Treatment consisted of surgery and adjuvant radiation or endocrine therapy. **(b)** Comparison of breast cancer PRS score distribution between patients with (red) or without (black) a breast cancer subsequent event (BCSE). Dashed vertical lines represent mean normalized PRS values for each respective group. Groups were compared with two-sided Mann-Whitney U test, and FDR corrected q-values were computed using Bonferroni corrected p-values for the effective number of tests. Distributions were generated using kernel density estimates of histograms. **(c)** Forest plot representation of hazard ratios (square) and 95% confidence intervals (error-bars), for each tested breast cancer PRS, obtained from a Cox Proportional-Hazard model accounting for DCIS size, grade, and age, the ancestry of the patient (Figure S4). The dotted line represents a log hazard ratio of 1, or having no effect on the outcome. The q-values represent Bonferroni corrected p-values for the effective number of tests. Significant hazard ratios (q<0.05) are indicated in bold text. **(d)** Evaluation of discrimination of Cox proportional hazard model for BCSE vs no BCSE outcome using Harrel’s C-index (y-axis) for models only using available risk factors versus available risk factors and breast cancer PRS, colored by the significance of hazard ratios for breast PRS (q<0.05, light green). **(e)** Same as (c) but for non-cancer PRS. **(f)** Cox proportional hazard estimate of breast cancer subsequent event (BCSE) - free survival for two overall and ER+ breast cancer PRS over time in years. Curves are obtained by varying PRS (solid colored lines from blue as lowest and red as highest PRS percentile), as compared to each model baseline (dashed line) while keeping all other covariates the same. Each case and control was weighted by the epidemiological incidence of BCSE treated with surgery and endocrine therapy (15% at 10 years) [24].

In order to evaluate the relationship between breast cancer PRS and DCIS prognosis, we curated 6 previously established breast cancer PRS, measuring risk for both overall and ER+ breast cancer, consisting of 859 total and 674 unique sites (see Methods, Table S2) [22,29,32–34]. We computed PRS for each patient, and compared groups with and without BCSE (Figure 3b) (see Methods). Patients with BCSE showed near significant elevated values across all 6 PRS (mean 1.4x fold increase, minimum p=0.06). We next measured the prognostic value of PRS in a multivariate Cox proportional hazard model to account for other risk factors previously associated with DCIS progression such as age, DCIS size, histological grade, and ancestry. We found that three of the breast cancer PRS had a significant (q<0.01) impact on BCSE risk, with the most impactful overall and ER+ breast cancer PRS hazard ratios of 2.5 (95%CI: 1.4–4.5, q=0.008) and 2.01 (1.3–3.1, q=0.007) respectively (Figure 3c, Figure S4). Adding PRS to the model improved the discrimination between patients with and without BCSE raising the mean C-index from 0.66 to 0.71 (Figure 3d). In contrast, none of the six non-cancer PRS contributed significantly to the BCSE prognosis, indicating that the effects observed are likely specific to the underlying genetic risk specific to breast cancer (Figure 3e). We estimate that 10 years post-DCIS diagnosis, approximately 24% of patients with the highest decile of breast PRS will have a BCSE, as opposed to approximately 3% of patients with PRS in the lowest decile (Figure 3f). Even with this limited dataset, there is a suggestive contribution of pre-established breast cancer PRS in DCIS prognosis, though this will require validation in a larger independent cohort. Independent of its possible clinical significance, and acknowledging the need for additional validation of the results, the presented use case demonstrates the feasibility of using DNA from tissues archived for decades to associate germline genetic factors with long-term patient outcomes and gain new insight into disease etiology and progression.

### Imputation of HLA-gene alleles from lc-WGS

In addition to SNP genotyping, we next investigated whether lc-WGS of archival tissue could be used to determine the haplotypes of the various HLA genes. HLA genes are some of the most polymorphic genes in the human genome and the major histocompatibility complex plays a critical role in antigen presentation to the immune system, particularly in tumorigenesis [35–37]. Using samples collected from 14 patients, including 10 patients with both blood and tissue DNA available, we compared HLA-alleles imputed from genome-wide genotypes obtained from lc-WGS against the results of clinical-grade deep targeted sequencing of the HLA locus from matching blood DNA samples (referred to as gold standard - see Methods). Alleles for Class I (HLA-A, B, C genes) and Class II (DRB1, DQB1 genes) were imputed using QUILT-HLA against the 1000G reference panel [6]. Overall 4 field allele calls from blood DNA were 92.8% (78/84) and 80.4% (45/56) concordant with the gold standard for Class I and Class II genes respectively (Figure 4a). At a lower 2 field resolution, the concordance was 97% for Class I and 91% for Class II (Figure S5). The decreased accuracy for HLA Class II, particularly for DRB1 likely reflects the increased diversity of these loci in comparison to Class I as well as the presence of pseudo-genes which may introduce ambiguity in the alignment of short sequence reads [38].

**Figure 4.**
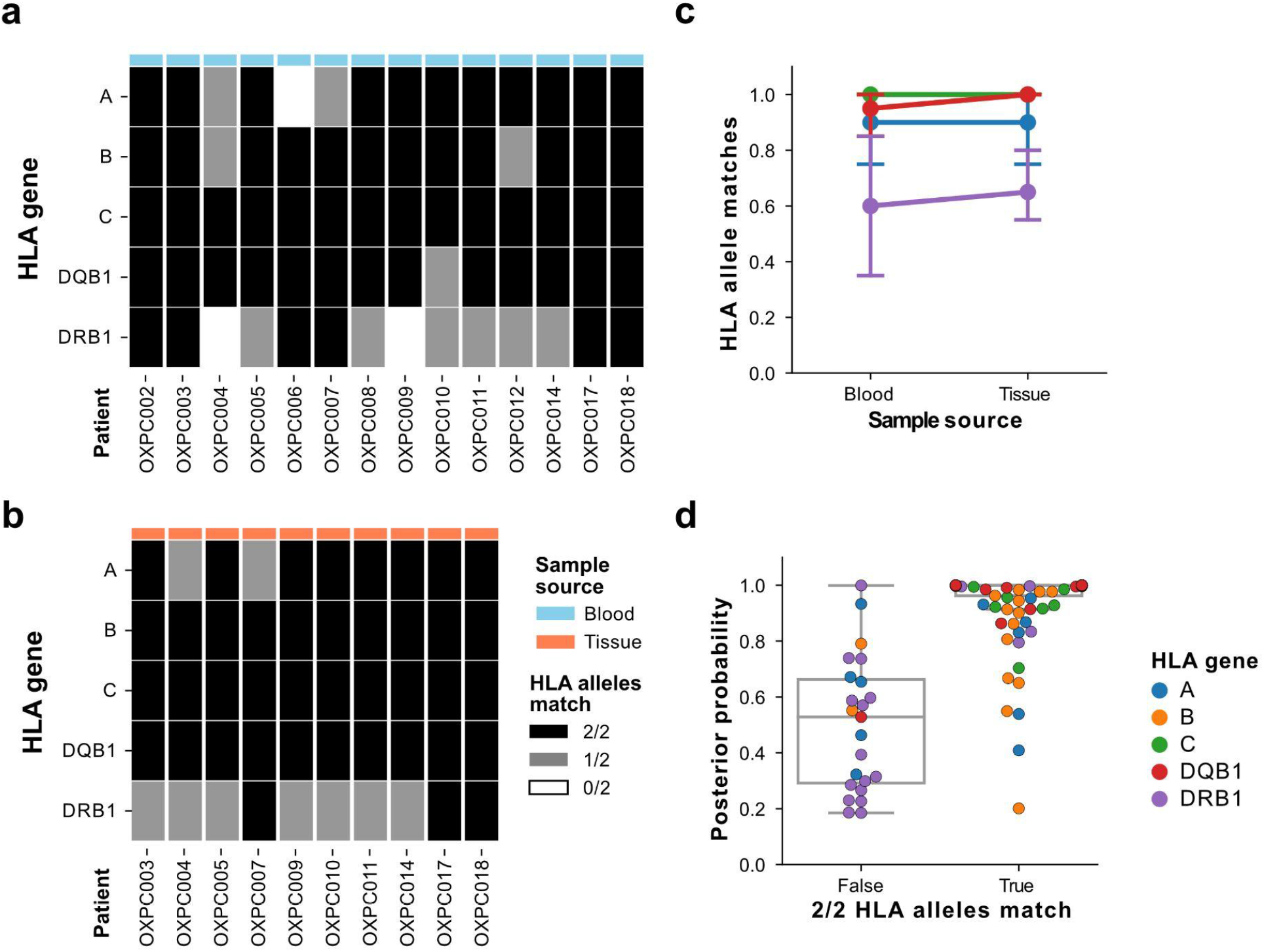
Assessment of 4 field HLA-typing accuracy from lc-WGS. **(a**,**b)** Number of concordant HLA alleles (0: white, 1: grey:, 2: black) between haplotypes from the clinical gold standard and those imputed using QUILT-HLA for class I (A, B, C) and class II (DQB1 and DRB1) HLA genes (rows) using (a) blood DNA of 14 patients or (b) tissue DNA of 10 patients (columns). **(c)** Fraction of HLA alleles correctly imputed (y-axis), versus the sample source of the DNA (x-axis), colored by the HLA gene. **(d)** Imputation posterior probability from QUILT-HLA for each HLA gene (color) and sample (dot), compared between samples with perfect HLA-gene concordance (both alleles match) versus those with errors.

In order to evaluate the effect of DNA source on HLA-typing accuracy, we compared tissue-derived HLA types to the gold standard. We found 4 field allele calls from tissue were 96.7% (58/60) and 82.5% (33/40) concordant with the gold standard blood HLA-typing, for Class I and Class II respectively (Figure 4b). In 49/50 comparisons between blood and tissue, tissue-derived samples provided as accurate calls, suggesting that the DNA source did not have an impact on imputation quality (Figure 4c). Overall, HLA-types that did not match the gold standard had worse imputation quality as reflected by their lower posterior probabilities (Figure 4d). The high accuracy of HLA-typing from lc-WGS as well as the consistent results between blood and tissue-based DNA demonstrates that remarkably, imputed HLA-types from lc-WGS on archival tissue are comparable against deep targeted HLA sequencing on blood, with a fraction of the required DNA input and a streamlined protocol.

## Discussion

Here we rethink the traditional design of germline genetic studies by answering the question, when typical DNA sources such as blood, saliva or urine are unavailable, can we extract the same information from archival tissue specimens? Often collected for histological examination and diagnosis and then stored indefinitely, these samples offer an abundant source of genetic material from patients with potentially long clinical follow-up. By using lc-WGS and recent advances in genotype imputation, we compared the concordance of germline genotypes obtained from blood DNA and archival tissue DNA in 10 different individuals. Archival tissue faithfully represented the germline profile of common SNPs obtained from blood both at the genome-wide level and across well-established PRS. Beyond concordance at the SNP-level, we also demonstrated accurate genotyping at highly polymorphic HLA alleles. To our knowledge, we present the first evidence that HLA-typing using lc-WGS from archival tissue is as accurate as true clinical-grade HLA-typing. Our results support the future utilization of archival tissue to construct large retrospective studies to characterize the role of germline variants in disease etiology, progression, and treatment.

The use of archival tissue as a source of constitutive DNA will enable a wealth of retrospective studies by repurposing specimens archived by most clinical sites to help address the genetic underpinnings of disease with long-outcome, such as the progression of pre-malignant lesions as presented here. Such studies would either require long follow up after the initial sample collection, or a massive and costly effort to retrospectively collect blood or saliva samples. In contrast, provided the subjects have been offered diagnostic biopsies, or surgical treatment, the course of their clinical care or study participation, their left-over specimen can be used to enable post-hoc genetic analysis. Of course, such studies would require approval of the Institutional Review Boards (IRB) and, since 2015, informed consent needs to be explicit about the use of specimens and data for genetic research and the risk for privacy it entails [39]. Commonly, IRBs waive the requirement for consent from patients deceased or lost to follow-up, however, such data needs to be distributed with caution and typically protected by a Data Access Policies the researcher has to comply with. As such, while our approach can enable large retrospective genetic studies where informed consent may be waived, the eligibility of each patient, and the overall data sharing policy need to be carefully considered.

Our report includes the application of the approach to interrogate the contribution of genetic factors to breast DCIS progression. The relatively good outcome of the disease poorly justified a thorough collection of risk variables, especially those related to inherited risk. However, overtreatment of DCIS, and its harms, is being increasingly acknowledged and systematic reviews of clinicopathological factors have not resulted in reliable models of progression [11,12,40]. Most epidemiological studies need to be large due to the slow progression and rarity of poor outcomes and rely exclusively on medical chart review [24,41,42]. As such, additional factors that are hard or impossible to collect from the charts such as mammography or magnetic resonance imaging, digital pathology, or germline inherited factors have not been as thoroughly and systematically investigated. We made the narrow hypothesis that lifetime breast cancer susceptibility - which can be seen as progression from normal to malignant epithelium - and progression of DCIS share the same genetic risk factors. We tested this hypothesis by measuring breast PRS in a small cohort of carefully selected DCIS subjects using our approach. Given the effect size of PRS contribution to breast cancer (HR=1.61), we anticipated that a balanced cohort of 36 patients would be sufficient to measure an effect size of HR=1.6 or greater representative of the contribution of other risk factors to DCIS progression [22,43,44]. Thanks to the accurate PRS estimate obtained from left-over surgical specimens, we were able to see that germline variation likely contributed significantly to the DCIS progression to an extent similar or greater to previously investigated risk factors such as grade, age, and Her2 overexpression [40]. Such findings would clearly need to be validated in a larger cohort, where a more comprehensive set of covariates would be accounted for, including treatment. Subsequent larger studies would also be important to evaluate competing risk models for subsequent in situ versus invasive disease, or laterality of the event, where PRS may contribute more in particular contexts. The modest cost and relative experimental simplicity of our approach, accompanied by a state-of-the-art imputation strategy can likely be scaled up provided diagnostic sections or left-over specimens can be found. Several large DCIS cohorts are generating mutational profiles, including some with lc-WGS and associated with clinical outcomes, which would be particularly suitable for validation in the future [17,45].

In the study of malignant progression as well as the onset and progression of multiple other diseases, the overactivity or inactivity of the immune system represents a key factor. A large contribution of variation in immune traits is inherited and yet the role of this contribution in disease progression is poorly understood [46,47]. In particular, the genetic diversity of the MHC, one of the most polymorphic regions of the genome, is a real challenge to study the role of the adaptive immune system. In the context of tumorigenesis, the failure of the major histocompatibility complex (MHC) to present antigens to the immune system is being increasingly recognized as contributing to cancer immune evasion and failure to respond to immune checkpoint inhibitors [48–50]. The determination of the HLA haplotypes, encoding the MHC is typically limited to the setting of organ or bone marrow transplants and not typically performed in other epidemiological studies. Recent reports however show the importance of the HLA-type in understanding the exposed mutanome, and its consideration can have important predictive value in the context of immunotherapies [35,36,51]. But with a lack of systemic HLA-typing or absence of genetic material to do so, such studies are hard to replicate or scale-up. To address this, we demonstrated that we can assign 4 field alleles to HLA-A, B, C, and DRB1, DQB1 genes by reference informed imputation of lc-WGS data [6]. These imputed HLA-types had comparable accuracy to deep targeted sequencing of the HLA locus with a fraction of the required DNA input (5 vs 40,000 ng) and with a simplified protocol (no need for targeted capture). The improvement in both sample requirement and throughput to HLA-typing supports the evaluation in lc-WGS with imputation in replacing current clinical standard tests.

While offering many benefits, there are still some limitations to lc-WGS paired with imputation for germline profiling of archival tissue. Similar to previous reports benchmarking lc-WGS imputation, error increases with decreasing minor allele frequency [5,6]. This would preclude the use of this strategy for the identification of rare variants of high penetrance associated with familiar risk (*BRCA*, Lynch, or Li-Fraumeni syndromes). Similarly, genotypes from rare risk-associated SNPs or HLA-types only found in small populations would be more likely missed by this approach. In the future, the availability of even larger and more diverse reference populations may help mitigate this effect. For the purposes of this study we utilized the unrestricted 1000G reference panel (N=3,202 haplotypes), however larger extensive, though restricted, panels such as Haplotype Reference Consortium (HRC) (N=64,976) or TopMed (N=53,831) exist [25,52,53]. Low coverage depth represents an additional limitation of our approach. While a restricted number of reads sequenced from a WGS library can result in decreased imputation accuracy, another source of tumor-specific decreased coverage is somatic copy number alterations (CNA). We observed that regions in a copy number loss resulted in decreased imputation accuracy. Similar observations were recently reported in a study performing germline imputation from discarded reads from targeted-sequencing tumor-derived tissue [1]. Here the choice of the tissue source, or the possibility to dissect normal histological regions, can help mitigate these effects. Indeed the use of adjacent normal tissue, pre-malignant or low-grade lesions or even lymphocytic aggregates, or lymph node specimens would enrich for diploid cells resulting in fewer inaccurate genomic regions. In contrast, imputation in high-grade lesions or invasive tumors with prominent aneuploidy needs to be carefully considered and may be mitigated in the largest dataset where available CNA profiles could be used as prior information in the imputation strategy.

## Conclusion

In conclusion, our study demonstrates that archival tumor tissue is an appropriate DNA source to measure germline genetic variation in lieu of normal tissue or blood. By shallow sequencing of the genome, and imputing missing sequences using haplotypes from thousands of individuals, the resulting genotypes, particularly for common SNPs and HLA alleles between blood and archival tissue were quite comparable. Especially in the study of slow progressing or rare diseases which may have been logistically unrealistic due to a long time to events and large sample numbers required, this framework has the potential to enable very large retrospective genetic studies, driving both basic research and translational discoveries.

## Materials and Methods

### Patient selection

For the tissue-blood benchmarking study, a total of N=14 Lung adenocarcinoma cancer patients with available tumor tissue and matching buffy coat in N=10 were selected from the Moores Cancer Center Tissue and Technology Shared Resource (BTTSR).

For the DCIS PRS study, a total of 50 patients were originally selected from the UC San Diego ATHENA DCIS registry - a retrospective registry approved by the UCSD and UCSF IRB. Case patients with BCSE were first selected on the basis of time to BCSE, surgery type, care location, and availability of archival tissue blocks. Control patients were then selected from patients without BCSE, with long follow-up time and matching cases for risk factors including age at DCIS, ancestry, DCIS grade, DCIS size, treatment type, ER, and Her2 status when available (Table 1).

### Sample Preparation

Blood DNA was extracted from 50 µL of buffy-coat using DNAeasy blood and tissue kit (Qiagen). Tissue blocks were sectioned in 5 µm scrolls and 3 to 5 scrolls were used to extract DNA with Covaris FFPE truXTRAC FFPE tNA kit using M220 Covaris Focused UltraSonicator (Covaris). DNA was quantified using Qubit dsDNA HS Assay Kit (Thermo Fisher Scientific).

### Low-coverage whole genome sequencing (lc-WGS)

Between 5-300ng of DNA was used as input for the library preparation using NEB Ultra II FS library preparation kit (New England Biolabs), which combines enzymatic fragmentation with end-repair and A-tailing in the same tube. Ligated and purified libraries were amplified using KAPA HiFi HotStart Real-time PCR 2X Master Mix (KAPA Biosystems). Samples were amplified with 5 μL of KAPA P5 and KAPA P7 primers. The reactions were denatured for 45 seconds (sec) at 98 °C and amplified 13–15 cycles for 15 sec at 98 °C, for 30 sec at 65 °C, and for 30 sec at 72 °C, followed by final extension for 1 min at 72 °C. Samples were amplified until they reached Fluorescent Standard 3, cycles being dependent on input DNA quantity and quality. PCR reactions were then purified using 1x AMPure XP bead clean-up and eluted into 20 μL of nuclease-free water. The amplified and purified libraries were analyzed using the Agilent 4200 Tapestation (D1000 ScreenTape) and quantified by fluorescence (Qubit dsDNA HS assay). Sample libraries with distinct indices were pooled in equimolar amounts, then sequenced to a target coverage of 0.5x, using paired-end 2×100bp reads on a NovaSeq 6000 (Illumina).

### Sequencing read processing and sample quality control

Sequencing libraries were deconvoluted using bcl2fastq [54]. Adapter sequences were trimmed from the raw fastq files using atropos (v1.1.31) [55]. The trimmed reads were then aligned to GRCh38 using bwa-mem (v0.7.17) [56]. Duplicate reads were then marked using biobambam (v2.0.87)[57]. Overall genome-wide coverage was measured using mosdepth (v0.2.6), and contamination was measured using verifyBamID2 (v1.0.6) [58,59]. For the DCIS, samples with less than 0.45x coverage or were estimated to be >5% contaminated were removed from downstream analyses.

### Imputation of genotypes from lc-WGS

Genome-wide genotypes were imputed using lc-WGS specific method GLIMPSE (v1.1.1) with the hg38 version of 1000G 30x NYGC reference panel (N=3,202 individuals) [5,25] Phasing and imputation were performed directly on BAM files in individual chunks of each chromosome using “GLIMPSE_phase”, and then the imputed variants were subsequently ligated together for each chromosome using “GLIMPSE_ligate”. We note that short insertions and deletions were excluded from any analysis as these are currently unreliable from lc-WGS and not currently imputed by the strategy implemented [1].

### Measuring imputation concordance

Imputation concordance between samples was summarized using squared Pearson correlation values obtained from the bcftools “stats” function (v1.9), which captures the correlation between allele dosages of variants in each minor allele frequency (MAF) bin [60]. Variants across all the autosomes were used in genome-wide benchmarking performance and all chromosomes for PRS evaluations.

### Copy number analysis

Copy number alterations (CNAs) were called using CNVkit (v0.9.9) in “wgs” mode, average bin size was set at 100,000 bp [61]. A set of unrelated normal tissues sequenced with the same protocol were used to generate a panel of normals used during CNA calling. Any bins with a log_2_ copy ratio lower than -15, were considered artifacts and removed. Breakpoints between copy number segments were determined using the circular binary segmentation algorithm (p < 10−4). Copy number genomic burden was computed as the sum of sizes of segments in a gain (log_2_(ratio) > 0.3) or loss (log_2_(ratio) < − 0.3) over the sum of the sizes of all segments.

### Clinical standard HLA genotyping

Reference HLA genotyping was performed on approximately 40 μg genomic DNA extracted from buffy-coat aliquots. Samples were prepared using targeted hybrid-capture with AlloSeq Tx17 reagents (CareDx). Samples were pooled and sequenced in 2×150 bp read-length on iSeq 100 instruments (Illumina). Sequence data was analyzed using Assign (v1.0.2) software (CareDx) and IMGT-HLA reference database (v3.43.0.1) [62].

### Measuring PRS

Polygenic risk scores (PRS) were computed using the following equation:

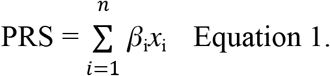

Equation 1. PRS is computed as a function of *β*_i_ which is the per-allele log odds ratio, or beta coefficient for the risk SNP allele i, and *x*_i_ is the dosage of the risk allele i {0,1,2}, and n is the total number of SNPs composing the PRS. PRS scores were then scaled using z-score transformation. PRS sites and effect weights were all obtained from the Polygenic Score (PGS) Catalog [63]. The catalog numbers and descriptions of each PRS are listed in Table S2.

### Cox proportional hazard model construction for breast PRS

Cox proportional hazard models were constructed non-parametrically, using Breslow’s method with robust estimates in lifelines survival analysis package in Python [64]. A separate model was constructed for each of the six evaluated breast PRS, in order to measure the effect on risk of BCSE, by PRS, DCIS nuclear grade, age of the patient at diagnosis, the size of the lesion, and whether the ancestry of the individual was European or not. Each covariate was tested for violation of the proportional hazards assumption. The 5 samples missing lesion size, were excluded from the model. In the 6 DCIS samples missing grade, grade was assigned on the basis of the tertiles of copy number burden distribution observed in the cohort since grade and copy number burden are highly correlated [14].

### Multiple hypothesis correction for non-independent PRS

In order to perform multiple hypothesis correction on multiple non-independent PRS, such as the breast PRS, we implemented the Li and Ji method in R package *meff* to estimate for the effective number of tests performed [65,66]. The effective number of tests was then used to generate Bonferroni corrected p-values, labeled as q-values.

### Ethics approval and consent to participate

The lung cancer patients were consented to the UC San Diego Biorepository and Tissue Technology Shared Resource, as a biorepository approved by the UC San Diego Institutional Review Board (protocols 090401 and 181755). The breast precancer patients were included in the ATHENA DCIS registry, a retrospective study approved by the UC San Diego Institutional Review Board (protocol 171481), which provided a waiver of consent.

### Consent for publication

Not applicable.

## Data Availability

Raw sequencing data pending deposition in dbGAP.

## Availability of data and materials

Raw sequencing data pending deposition in dbGAP.

## Competing interests

O.H. is an employee of Zentalis Pharmaceuticals.

## Funding

This work is supported by funding from the National Institute of Health (U01CA196406, U01CA196406, U01CA196383, T32GM008806, T15LM011271), and an award from the Cancer Cell Mapping Initiative (U54 CA209891), shared resources from the National Cancer Institute Cancer Center Support Grant (P30CA023100), The Mark Foundation for Cancer Research grant #18-022-ELA to HC and the California Tobacco-Related Disease Research Program pre-doctoral fellowship to DN (28DT-0011). The funding bodies had no role in the design of the study; collection, analysis, and interpretation of data; or in the writing of the manuscript.

## Authors’ contributions

D.N., L.B., M.P. performed the analysis, J.S., C.C., G.P.M., D.N. generated the data, N.Q.L., T.J.O., G.Y.L., F.H. collected the specimen and reviewed the clinical data. O.H., H.C. directed the study. O.H, D.N. wrote the manuscript. All authors reviewed and approved the manuscript.

## Acknowledgments

We thank Adam Maihofer for his statistical advice, Sharmeela Kaushal, Valeria Estrada, Kimberly Mcintyre, and the staff of the Moores Cancer Center Biorepository and Tissue Technology Shared Resources for the samples collection and processing. We are grateful to Kristen Jepsen and Huazhen Yao from the IGM genomics center for their technical expertise and sample genotyping and sequencing. We also kindly thank CareDx for providing the reagents for the targeted HLA sequencing.

## Supplemental Tables and Figures

**Figure S1.**
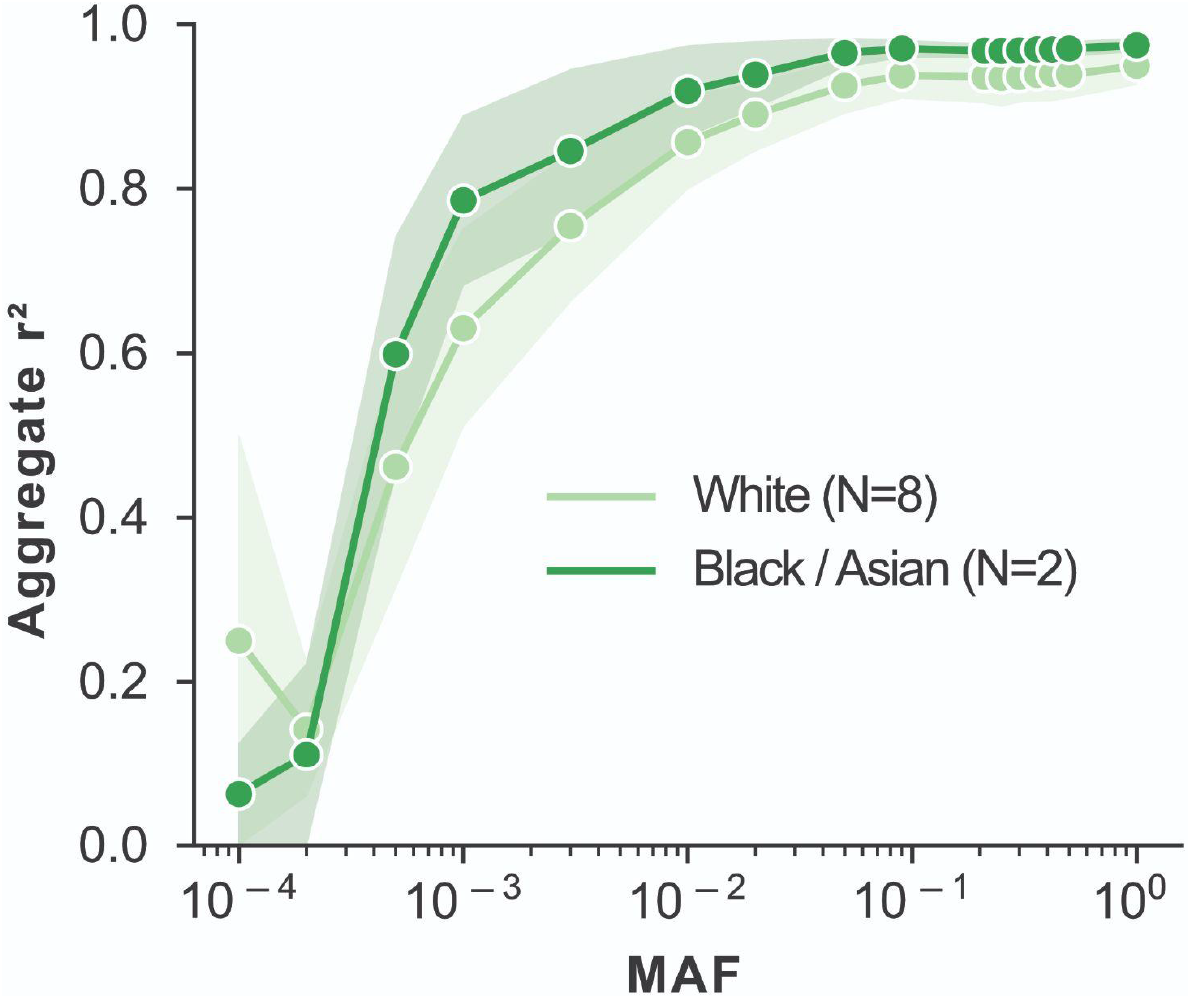
Effect of patient ancestry on lc-WGS imputation concordance between blood and tissue. Comparison of concordance between blood and tissue-based on ancestry background of the patient, with White ancestry in light green and Black or Asian ancestry in dark green. Pearson correlation squared (r^2^) is for all aggregated SNPs within a MAF bin. When available, 95% confidence intervals are shaded around the line based on 1000 bootstrap iterations.

**Figure S2.**
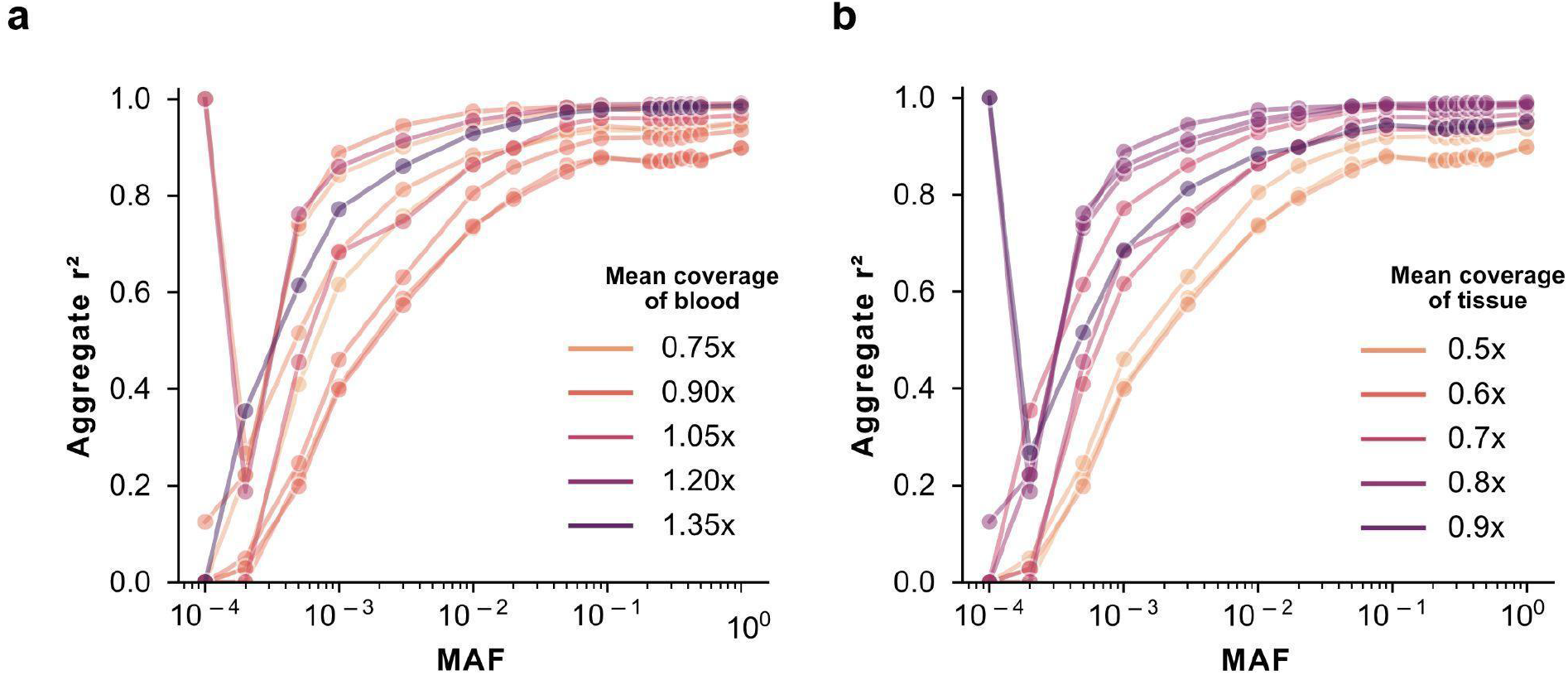
Effect of coverage-related features on lc-WGS imputation concordance between blood and tissue. **(a**,**b)** Comparison of concordance as measured by squared Pearson correlation (y-axis) between blood and tissue as a function of MAF (x-axis) based on mean sequencing genome coverage depth of blood (a) or tissue (b).

**Figure S3.**
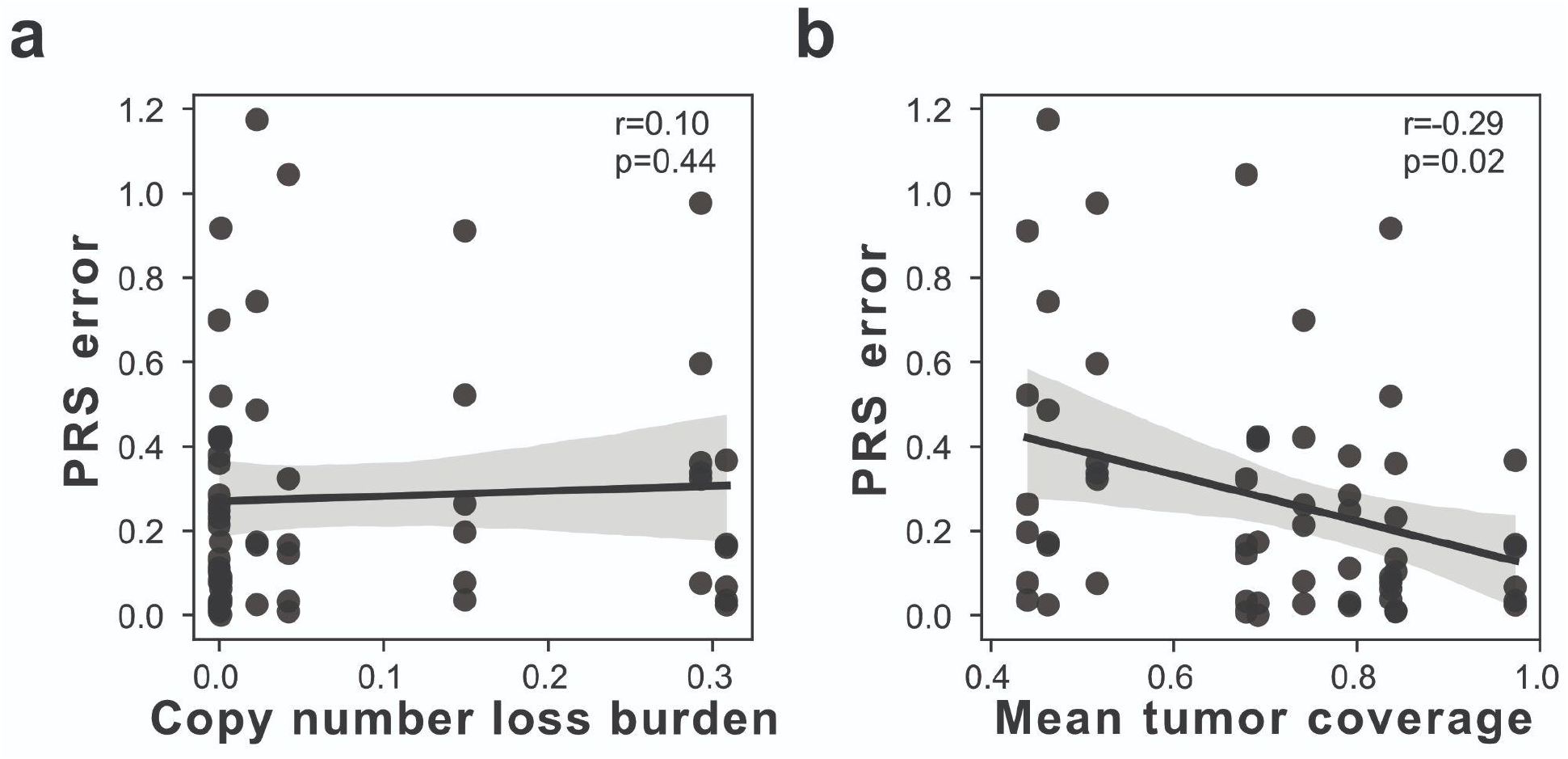
Effect of coverage-related features on the error in non-cancer PRS calculation. (a-b) PRS error (y-axis), as measured by the absolute difference between blood and tissue PRS across all non-cancer PRS, as a function of (a) fraction of genome in a copy number loss or (b) mean tissue/tumor genome coverage (x-axis). The 95% confidence intervals are shaded around the line based on 1000 bootstrap iterations. Spearman correlation coefficient, r, and corresponding p-value are indicated as text in the upper right.

**Figure S4.**
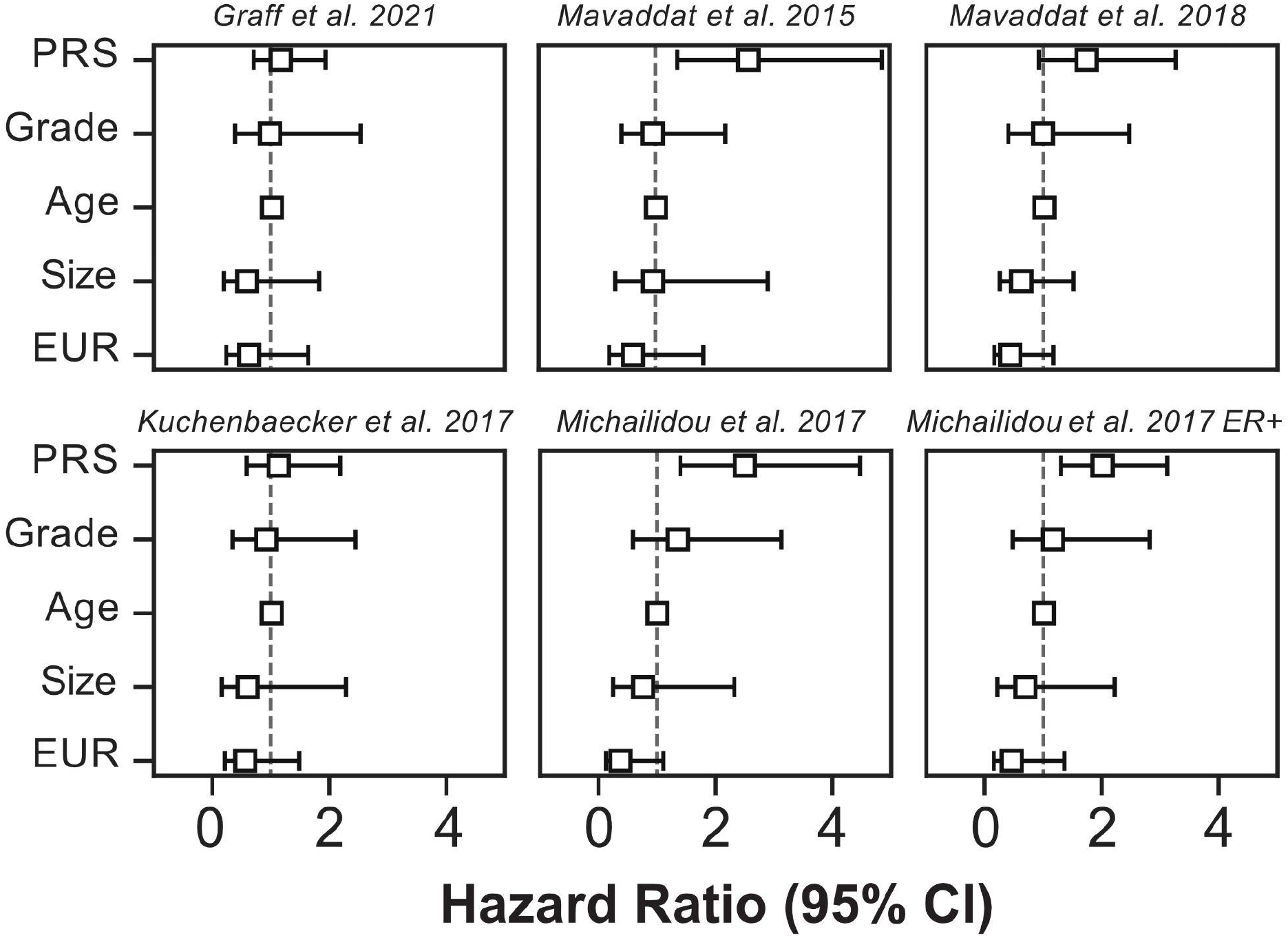
Cox proportional hazard models measuring BCSE outcome in DCIS patients for 6 breast cancer PRS. Forest plot representation of hazard ratios (square) and 95% confidence intervals (error-bars), for each normalized breast cancer PRS and covariates for DCIS BCSE risk including DCIS nuclear grade (Grade), age of the patient at diagnosis (Age), the size of the DCIS lesion (Size), and whether the ancestry of the individual was European (EUR). The dotted line represents a hazard ratio of 1, indicating no effect on BCSE risk, >1 indicating increased, and <1 indicating decreased risk.

**Figure S5.**
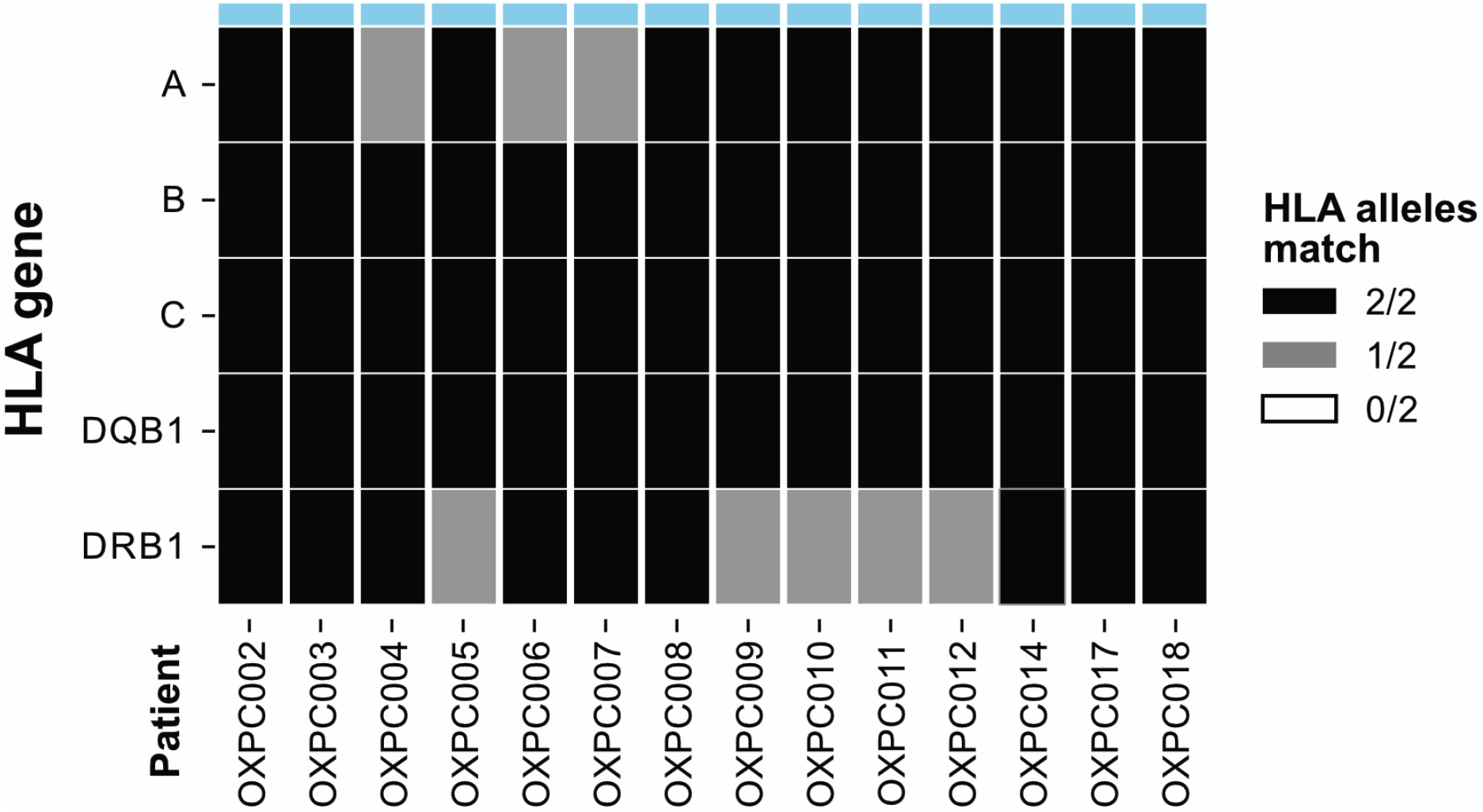
Assessment of 2 field HLA-typing accuracy from lc-WGS. Number of concordant HLA alleles (0: white, 1: grey:, 2: black) between haplotypes from the clinical gold standard and those imputed using QUILT-HLA for class I (A, B, C) and class II (DQB1 and DRB1) HLA genes (rows) using blood DNA of 14 patients.

**Table S1.** Description of the studied samples.

| **Patient** | **DNA source** | **Ancestry** | **Input DNA (ng)** | **Coverage** | **CNA burden** | **Age of block (yr)<sup>1</sup>** | **Analysis<sup>2</sup>** |
| --- | --- | --- | --- | --- | --- | --- | --- |
| OXPC002 | Blood | White | 68.4 | 1.11 | 0.00 | - | H |
| OXPC003 | Blood | White | 31.8 | 1.41 | 0.00 | - | GPH |
| OXPC003 | Tissue | White | 28.4 | 0.69 | 0.00 | 7 | GPH |
| OXPC004 | Blood | White | 32.1 | 0.84 | 0.00 | - | GPH |
| OXPC004 | Tissue | White | 5.5 | 0.44 | 0.15 | 7 | GPH |
| OXPC005 | Blood | White | 53.1 | 0.79 | 0.00 | 5 | GPH |
| OXPC005 | Tissue | White | 21.8 | 0.97 | 0.48 | - | GPH |
| OXPC006 | Blood | White | 48.9 | 0.95 | 0.00 | - | H |
| OXPC007 | Blood | White | 54.6 | 0.87 | 0.00 | - | GPH |
| OXPC007 | Tissue | White | 35.1 | 0.46 | 0.08 | 6 | GPH |
| OXPC008 | Blood | White | 45.3 | 0.89 | 0.00 | - | H |
| OXPC009 | Blood | White | 12.2 | 1.10 | 0.00 | - | GPH |
| OXPC009 | Tissue | White | 11.0 | 0.84 | 0.01 | 5 | GPH |
| OXPC010 | Blood | Black or African American | 16.9 | 0.85 | 0.00 | - | GPH |
| OXPC010 | Tissue | Black or African American | 176.4 | 0.84 | 0.01 | 5 | GPH |
| OXPC011 | Blood | White | 53.4 | 0.89 | 0.00 | - | GPH |
| OXPC011 | Tissue | White | 20.4 | 0.52 | 0.65 | 5 | GPH |
| OXPC012 | Blood | White | 66.0 | 0.71 | 0.00 | - | H |
| OXPC014 | Blood | White | 12.1 | 0.68 | 0.00 | - | GPH |
| OXPC014 | Tissue | White | 15.0 | 0.68 | 0.11 | 3 | GPH |
| OXPC017 | Blood | White | 24.2 | 0.71 | 0.00 | - | GPH |
| OXPC017 | Tissue | White | 30.6 | 0.79 | 0.01 | 3 | GPH |
| OXPC018 | Blood | Asian | 14.8 | 1.01 | 0.00 | - | GPH |
| OXPC018 | Tissue | Asian | 25.3 | 0.74 | 0.00 | 8 | GPH |

1. Age of block inferred as the difference in years from DNA extraction (2021) to the year of the diagnosis. 2. Analysis type sample was used in, which was one of: GPH: Genome wide, PRS and HLA H: HLA only (for samples without both tissue and blood)

**Table S2.**
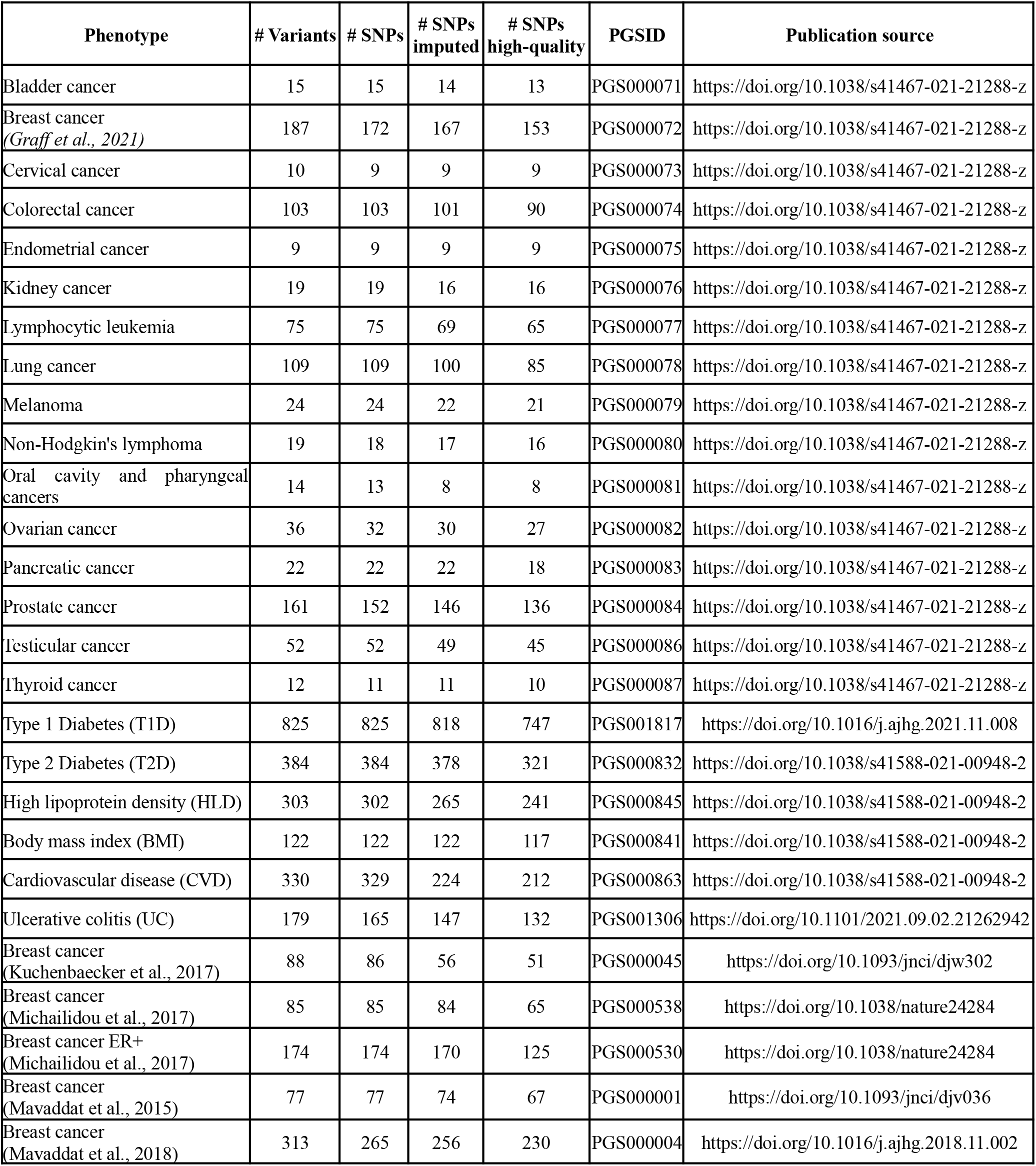
PRS description.

| Phenotype | # Variants | # SNPs | # SNPs imputed | # SNPs high-quality | PGSID | Publication source |
| --- | --- | --- | --- | --- | --- | --- |
| Bladder cancer | 15 | 15 | 14 | 13 | PGS000071 | https://doi.org/10.1038/s41467-021-21288-z |
| Breast cancer<br>(Graff et al., 2021) | 187 | 172 | 167 | 153 | PGS000072 | https://doi.org/10.1038/s41467-021-21288-z |
| Cervical cancer | 10 | 9 | 9 | 9 | PGS000073 | https://doi.org/10.1038/s41467-021-21288-z |
| Colorectal cancer | 103 | 103 | 101 | 90 | PGS000074 | https://doi.org/10.1038/s41467-021-21288-z |
| Endometrial cancer | 9 | 9 | 9 | 9 | PGS000075 | https://doi.org/10.1038/s41467-021-21288-z |
| Kidney cancer | 19 | 19 | 16 | 16 | PGS000076 | https://doi.org/10.1038/s41467-021-21288-z |
| Lymphocytic leukemia | 75 | 75 | 69 | 65 | PGS000077 | https://doi.org/10.1038/s41467-021-21288-z |
| Lung cancer | 109 | 109 | 100 | 85 | PGS000078 | https://doi.org/10.1038/s41467-021-21288-z |
| Melanoma | 24 | 24 | 22 | 21 | PGS000079 | https://doi.org/10.1038/s41467-021-21288-z |
| Non-Hodgkin's lymphoma | 19 | 18 | 17 | 16 | PGS000080 | https://doi.org/10.1038/s41467-021-21288-z |
| Oral cavity and pharyngeal cancers | 14 | 13 | 8 | 8 | PGS000081 | https://doi.org/10.1038/s41467-021-21288-z |
| Ovarian cancer | 36 | 32 | 30 | 27 | PGS000082 | https://doi.org/10.1038/s41467-021-21288-z |
| Pancreatic cancer | 22 | 22 | 22 | 18 | PGS000083 | https://doi.org/10.1038/s41467-021-21288-z |
| Prostate cancer | 161 | 152 | 146 | 136 | PGS000084 | https://doi.org/10.1038/s41467-021-21288-z |
| Testicular cancer | 52 | 52 | 49 | 45 | PGS000086 | https://doi.org/10.1038/s41467-021-21288-z |
| Thyroid cancer | 12 | 11 | 11 | 10 | PGS000087 | https://doi.org/10.1038/s41467-021-21288-z |
| Type 1 Diabetes (T1D) | 825 | 825 | 818 | 747 | PGS001817 | https://doi.org/10.1016/j.ajhg.2021.11.008 |
| Type 2 Diabetes (T2D) | 384 | 384 | 378 | 321 | PGS000832 | https://doi.org/10.1038/s41588-021-00948-2 |
| High lipoprotein density (HLD) | 303 | 302 | 265 | 241 | PGS000845 | https://doi.org/10.1038/s41588-021-00948-2 |
| Body mass index (BMI) | 122 | 122 | 122 | 117 | PGS000841 | https://doi.org/10.1038/s41588-021-00948-2 |
| Cardiovascular disease (CVD) | 330 | 329 | 224 | 212 | PGS000863 | https://doi.org/10.1038/s41588-021-00948-2 |
| Ulcerative colitis (UC) | 179 | 165 | 147 | 132 | PGS001306 | https://doi.org/10.1101/2021.09.02.21262942 |
| Breast cancer<br>(Kuchenbaecker et al., 2017) | 88 | 86 | 56 | 51 | PGS000045 | https://doi.org/10.1093/jnci/djw302 |
| Breast cancer<br>(Michailidou et al., 2017) | 85 | 85 | 84 | 65 | PGS000538 | https://doi.org/10.1038/nature24284 |
| Breast cancer ER+<br>(Michailidou et al., 2017) | 174 | 174 | 170 | 125 | PGS000530 | https://doi.org/10.1038/nature24284 |
| Breast cancer<br>(Mavaddat et al., 2015) | 77 | 77 | 74 | 67 | PGS000001 | https://doi.org/10.1093/jnci/djv036 |
| Breast cancer<br>(Mavaddat et al., 2018) | 313 | 265 | 256 | 230 | PGS000004 | https://doi.org/10.1016/j.ajhg.2018.11.002 |

**Table S3.**
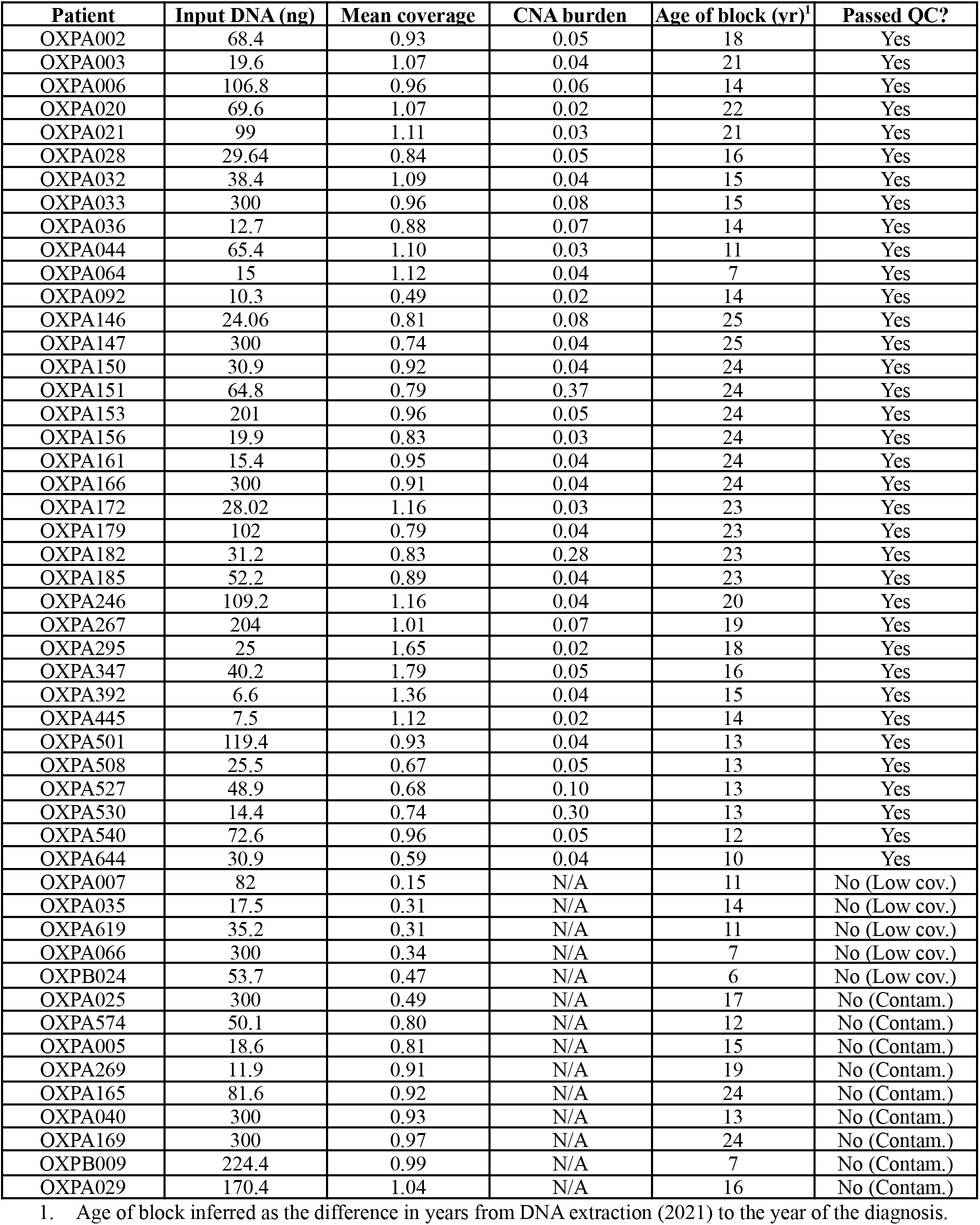
DCIS cohort technical characteristic description.

| Patient | Input DNA (ng) | Mean coverage | CNA burden | Age of block (yr)<sup>1</sup> | Passed QC? |
| --- | --- | --- | --- | --- | --- |
| OXPA002 | 68.4 | 0.93 | 0.05 | 18 | Yes |
| OXPA003 | 19.6 | 1.07 | 0.04 | 21 | Yes |
| OXPA006 | 106.8 | 0.96 | 0.06 | 14 | Yes |
| OXPA020 | 69.6 | 1.07 | 0.02 | 22 | Yes |
| OXPA021 | 99 | 1.11 | 0.03 | 21 | Yes |
| OXPA028 | 29.64 | 0.84 | 0.05 | 16 | Yes |
| OXPA032 | 38.4 | 1.09 | 0.04 | 15 | Yes |
| OXPA033 | 300 | 0.96 | 0.08 | 15 | Yes |
| OXPA036 | 12.7 | 0.88 | 0.07 | 14 | Yes |
| OXPA044 | 65.4 | 1.10 | 0.03 | 11 | Yes |
| OXPA064 | 15 | 1.12 | 0.04 | 7 | Yes |
| OXPA092 | 10.3 | 0.49 | 0.02 | 14 | Yes |
| OXPA146 | 24.06 | 0.81 | 0.08 | 25 | Yes |
| OXPA147 | 300 | 0.74 | 0.04 | 25 | Yes |
| OXPA150 | 30.9 | 0.92 | 0.04 | 24 | Yes |
| OXPA151 | 64.8 | 0.79 | 0.37 | 24 | Yes |
| OXPA153 | 201 | 0.96 | 0.05 | 24 | Yes |
| OXPA156 | 19.9 | 0.83 | 0.03 | 24 | Yes |
| OXPA161 | 15.4 | 0.95 | 0.04 | 24 | Yes |
| OXPA166 | 300 | 0.91 | 0.04 | 24 | Yes |
| OXPA172 | 28.02 | 1.16 | 0.03 | 23 | Yes |
| OXPA179 | 102 | 0.79 | 0.04 | 23 | Yes |
| OXPA182 | 31.2 | 0.83 | 0.28 | 23 | Yes |
| OXPA185 | 52.2 | 0.89 | 0.04 | 23 | Yes |
| OXPA246 | 109.2 | 1.16 | 0.04 | 20 | Yes |
| OXPA267 | 204 | 1.01 | 0.07 | 19 | Yes |
| OXPA295 | 25 | 1.65 | 0.02 | 18 | Yes |
| OXPA347 | 40.2 | 1.79 | 0.05 | 16 | Yes |
| OXPA392 | 6.6 | 1.36 | 0.04 | 15 | Yes |
| OXPA445 | 7.5 | 1.12 | 0.02 | 14 | Yes |
| OXPA501 | 119.4 | 0.93 | 0.04 | 13 | Yes |
| OXPA508 | 25.5 | 0.67 | 0.05 | 13 | Yes |
| OXPA527 | 48.9 | 0.68 | 0.10 | 13 | Yes |
| OXPA530 | 14.4 | 0.74 | 0.30 | 13 | Yes |
| OXPA540 | 72.6 | 0.96 | 0.05 | 12 | Yes |
| OXPA644 | 30.9 | 0.59 | 0.04 | 10 | Yes |
| OXPA007 | 82 | 0.15 | N/A | 11 | No (Low cov.) |
| OXPA035 | 17.5 | 0.31 | N/A | 14 | No (Low cov.) |
| OXPA619 | 35.2 | 0.31 | N/A | 11 | No (Low cov.) |
| OXPA066 | 300 | 0.34 | N/A | 7 | No (Low cov.) |
| OXPB024 | 53.7 | 0.47 | N/A | 6 | No (Low cov.) |
| OXPA025 | 300 | 0.49 | N/A | 17 | No (Contam.) |
| OXPA574 | 50.1 | 0.80 | N/A | 12 | No (Contam.) |
| OXPA005 | 18.6 | 0.81 | N/A | 15 | No (Contam.) |
| OXPA269 | 11.9 | 0.91 | N/A | 19 | No (Contam.) |
| OXPA165 | 81.6 | 0.92 | N/A | 24 | No (Contam.) |
| OXPA040 | 300 | 0.93 | N/A | 13 | No (Contam.) |
| OXPA169 | 300 | 0.97 | N/A | 24 | No (Contam.) |
| OXPB009 | 224.4 | 0.99 | N/A | 7 | No (Contam.) |
| OXPA029 | 170.4 | 1.04 | N/A | 16 | No (Contam.) |

1. Age of block inferred as the difference in years from DNA extraction (2021) to the year of the diagnosis.

**Table S4.** DCIS cohort covariate association with patient outcome.

| Clinical feature |  | No BCSE<sup>2</sup><br>(N=14) | BCSE<br>(N=22) | Significance<sup>3</sup> |
| --- | --- | --- | --- | --- |
| **Grade<sup>1</sup>** | Low | 36% (5) | 41% (9) | *p*=0.59 |
|  | Intermediate | 50% (7) | 27% (6) |  |
|  | High | 7% (1) | 9% (2) |  |
| **ER status<sup>1</sup>** | + | 71% (10) | 77% (17) | *p*=0.39 |
|  | - | 7% (1) | 0% (0) |  |
| **Ethnicity<sup>1</sup>** | Hispanic | 14% (2) | 9% (2) | *p*=0.63 |
|  | Non-Hispanic | 86% (12) | 91% (20) |  |
| **Race** | Asian | 14% (2) | 9% (2) | *p*=0.63 |
|  | White | 86% (12) | 91% (20) |  |
| **Pathologic size (cm)<sup>1</sup>** |  | 1.35 | 0.85 | *p*=0.05 |
| **Age at diagnosis (yrs)** |  | 56.3 | 60.1 | *p*=0.27 |

1. Missing values not represented here, but can be found in Table S2. 2. BCSE: Breast cancer subsequent event. 3. P-values were computed using Fisher Exact test for ER status, Race, Ethnicity and Chi-square test for Grade. For continuous features, size and age, Mann-Whitney U test was used to compare groups.

## Notes

### Competing Interest Statement

Dr Harismendy is a current employee of Zentalis Pharmaceuticals

